# Artificial neural network predicts sex differences of patients with advanced Parkinson’s disease under Levodopa-Carbidopa Intestinal gel

**DOI:** 10.1101/2023.06.26.23291833

**Authors:** Anastasia Bougea, Tajedin Derikvand, Efthymia Efthymiopoulou, Efthalia Angelopoulou

**Affiliations:** 1st Department of Neurology, Eginition Hospital, National and Kapodistrian University of Athens, Greece; Department of Mathematics, Marvdasht Branch, Islamic Azad University, Marvdasht, Iran; AbbVie Inc., Greece

**Keywords:** advanced Parkinson’s disease (PD), non-motor symptoms (NMS), Levodopa*-* carbidopa intestinal gel(LCIG), Non-Motor Symptoms Questionnaire (NMSQ), NMS Questionnaire (NMSQ), Geriatric Depression Scale (GDS)Unified Parkinson’s Disease Rating Scale (UPDRS), Long Short-Term Memory-recurrent neural network (LSTM-RNN)

## Abstract

**Objective:** Although Levodopa-carbidopa intestinal gel (LCIG) treatment has shown to be efficacious in motor and some non-motor symptoms (NMS), not all the patients with advanced Parkinson’s disease (PD) are ideal candidates. To improve their selection analysis knowledge of prognostic factors is of great importance. We aimed to develop a novel machine learning model to predict the clinical outcomes of patients with advanced PD at 2 years under the LCIG therapy.

**Methods:** This was a longitudinal 24-month, observational study of 59 patients with advanced PD of a Greek multicenter registry under LCIG treatment from September 2019 to September 2021. Motor status was assessed with the Unified Parkinson’s Disease Rating Scale (UPDRS) part III (off) and IV. NMS were assessed by the NMS Questionnaire (NMSQ) and the Geriatric Depression Scale (GDS), the quality of life by PDQ-39 and severity by Hoehn &Yahr (HY). Multivariate linear regression, ARIMA, SARIMA, and Long Short-Term Memory-recurrent neural network (LSTM-RNN) models were used.

**Results:** Dyskinesia duration and quality of life were significantly improved with LCIG (19% and 10% greater improvement for men than women, respectively). Multivariate linear regression models showed that UPDRS-III was decreased by 1.5 and 4.39 units per one unit of increase of the PDQ-39, UPDRS-IV indexes, respectively. Among all the time series models, the LSTM-RNN model predicts these clinical characteristics with highest accuracy (mean square error =0.0069)

**Conclusions:** Τhe LSTM-RNN model predicts with highest accuracy sex dependent clinical outcomes of patients with advanced PD after two years of LCIG therapy.

## Introduction

Parkinson’s disease (PD) is a chronic neurodegenerative disease characterized by a complex range of motor and non-motor symptoms with a major impact on the patient’s quality of life (QoL)^1, 2^. In the advanced stages of PD, patients may experience unsuccessful control of the motor complications through levodopa oral/transdermal medications. At this stage there are available several invasive therapeutic options (device-aided therapies) to be considered ^3^. This lack of control could be explained by the inability to buffer exogenous dopamine that results from the denervation in the striatum, the short half-life of Levodopa and the delayed gastric emptying which causes un predictable fluctuation of plasma Levodopa.

The continuous delivery of levodopa as levodopa-carbidopa intestinal gel (LCIG) may overcome the above limitations improving motor/non-motor symptoms of patients with advanced PD and their QoL^4^. However, not all the patients with advanced PD are ideal candidates. To improve their selection analysis, deep knowledge of prognostic factors is of great importance. Moreover prospective studies specifically addressing sex differences in LCIG treatments’ outcomes are lacking.

In this regard, artificial intelligence including machine learning models (ML) could assist in treatment optimization for advanced PD^5–8^. One of the most advanced ML models to forecast time series is the Long Short-Term Memory (LSTM) Neural Network^9^. Autoregressive Integrated Moving Average (ARIMA) models provide another approach to time series forecasting under the premise that past values have an effect on current values, which makes this statistical technique popular for analyzing nature, health, and other processes that vary over time. Moving average (MA) term in a time series model is a past error (multiplied by a coefficient). A seasonal autoregressive integrated moving average (SARIMA) model is one step different from an ARIMA model, based on the concept of seasonal trend ^10^.

The aim of this study was to develop the best ML neural network model by the comparison with the above models-to predict the progression of both motor and non-motor symptoms of patients at 2 years of follow-up after the implementation of LCIG.

## Methods

### Study Population and Design

ForHealth S.A is a Greek multicenter, observational registry for patients with advanced PD under LCIG therapy, including 43 movement disorder centers in greek cities (Athens, Thessaloniki, Patra, Ioannina, Chania, Larisa). This was a longitudinal 24-month, multicenter, non-interventional, observational study of 59 patients of ForHealth S.A with advanced PD under LCIG treatment from September 2019 to September 2021. Inclusion criteria were patients with advanced PD on stable LCIG regimens for the 24-month follow-up not receiving any other antiparkinsonian medication. Exclusion criteria included dementia and major depression. The following data were collected: demographics, disease duration, motor and non-motor clinical measures in years 0 and 2, and levodopa equivalent daily dose (LEDD).

LCIG (20 mg/mL) was administered via a percutaneous endoscopic gastrostomy tube with jejunal extension (PEG-J) over 16 hours using a portable pump (CADD-Legacy®, Duodopa CE 0473) ^4^. LCIG treatment was composed of 3 individually adjusted doses: the morning bolus dose (usually 5-10 mL [100-200 mg levodopa]); the continuous maintenance dose (usually 2-6 mL/hour [40-120 mg levodopa/hour]); and extra bolus doses, adjusted individually. Motor status was assessed with the Unified Parkinson’s Disease Rating Scale part III (UPDRS-III) in off stage. Hours of “Off” and dyskinesia duration were assessed with the Unified Parkinson’s Disease Rating Scale part IV (UPDRS-IV) ^11^. Non-motor symptoms were evaluated by the Non-Motor Symptoms Questionnaire (NMSQ) ^12^ and Geriatric Depression Scale (GDS) ^13^. The severity of PD was assessed using the Hoehn and Yahr (HY)^14^ and quality of life by PD questionnaire-39 (PDQ-39) ^15^. All procedures performed in studies involving human participants were in accordance with the ethical standards of the institutional and/or national research committee and with the 1964 Helsinki Declaration and its later amendments or comparable ethical standards. The study protocol was approved by ethics committee of AbbVie Greece approval no.:20-6-2019.

### Statistical Analysis

Patient’s demographics were summarized using descriptive statistics. Quantitative data were expressed as mean=:J±=:Jstandard deviation (SD). In data preprocessing phase, some missing data were imputed, using their column’s mean and one-dimensional wavelet transform was used to remove possible noise in all columns. Shapiro-Wilk and Kolmogorov Smirnov normality tests confirmed normal distribution of all features in men and women subgroups before and after LCIG treatment. Independent samples t-test and paired samples t-test were used to compare the LCIG effects on QL in PD patients. A multi-regression model was fitted for the measured clinical characteristics of men and women separately, and the possible synergistic effect of the features were evaluated. Irregular time series were turned into regular ones using the spline interpolation method. The best seasonal ARIMA model was derived for the time series, and clinical measures were forecasted for a horizon of the ten following time points. A ML method was presented to classify patients, and then the class or grade of the new patients could be determined based on their known PD characteristics. LSTM model was used to predict future values based on previous, sequential data. The R language and environment was used for statistical computing and graphics. Statistical significance was a set at p=0.05.

## Results

Fifty nine patients were included in the study. At baseline, mean age was 71 ± 10.18years, and47.6% were men. Mean disease duration was 14.28 ± 5.64 years. At baseline, clinical characteristics were the following: dyskinesia duration (2.44± 0.61), HY (2.05± 0.2), UPDRS-III (off) (30.50 ± 4.15), UPDRS-IV (3.13± 1.02), GDS (7.48± 2.42), NMSQ (10.09±0.97) and PDQ-39 (37.50 ±0.79). Main demographic and clinical characteristics are summarized in Table 1.

**Table 1.** Clinical features of patients with PD before and after using LCIG Treatment

|  | **Men**<br>**(Before LCIG**<br>**Treatment)** | **Men**<br>**(After LCIG**<br>**Treatment)** | **Women**<br>**(Before**<br>**LCIG**<br>**Treatment)** | **Women**<br>**(After LCIG**<br>**Treatment)** |
| --- | --- | --- | --- | --- |
| **Age** | **70 ± 12.08** | **-** | **73.45 ± 8.35** | **-** |
| **PD Duration** | **14.3 ± 4.83** | **-** | **14.27 ± 6.52** | **-** |
| **GDS** | **8.18 ± 2.09** | **-** | **6.7 ± 2.62** | **-** |
| **Dyskinesia Duration** | **1.86 ± 0.16** | **0.67 ± 0.17** | **3.01 ± 0.15** | **1.49 ± 0.18** |
| **“Off” Duration** | **5.26 ± 0.25** | **4.57 ± 0.43** | **5.94 ± 0.25** | **4.73 ± 0.52** |
| **NMSQ** | **9.15 ± 0.24** | **7.89 ± 0.84** | **11.01 ± 0.15** | **9.53 ± 1.27** |
| **UPDRS-III (state off)** | **32.55 ± 4.78** | **33.76 ± 3.43** | **28.45 ± 2.03** | **23.79 ± 4.78** |
| **UPDRS-IV** | **2.14 ± 0.12** | **2.34 ± 0.93** | **4.11 ± 0.25** | **3.90 ± 0.09** |
| **HY** | **2.21 ± 0.18** | **2.04 ± 0.08** | **1.88 ± 0.10** | **1.75 ± 0.20** |
| **PDQ-39** | **37.31 ± 0.94** | **30.77 ± 1.30** | **37.67 ± 0.59** | **29.61 ± 2.19** |

GDS, Geriatric Depression Scale; HY, Hoehn and Yahr; LCIG, Levodopa-carbidopa intestinal gel; NMSQ, Non-Motor Symptoms Questionnaire; PDQ, Parkinson's Disease; PDQ-39, Parkinson's Disease Questionnaire; UPDRS III, Unified Parkinson's Disease Rating Scale Part III

Our data set includes seven clinical features of PD patients before and after LCIG implementation (Table 1). Although, all features are improved in men and women, dyskinesia duration is decreased significantly in both groups and the variation of UPDRS-III is sex dependent.

LCIG significantly improves quality of life, but its effects on dyskinesia duration and UPDRS-III (off) score varies by sex (Figure 1 and 2).

**Figure 1.**
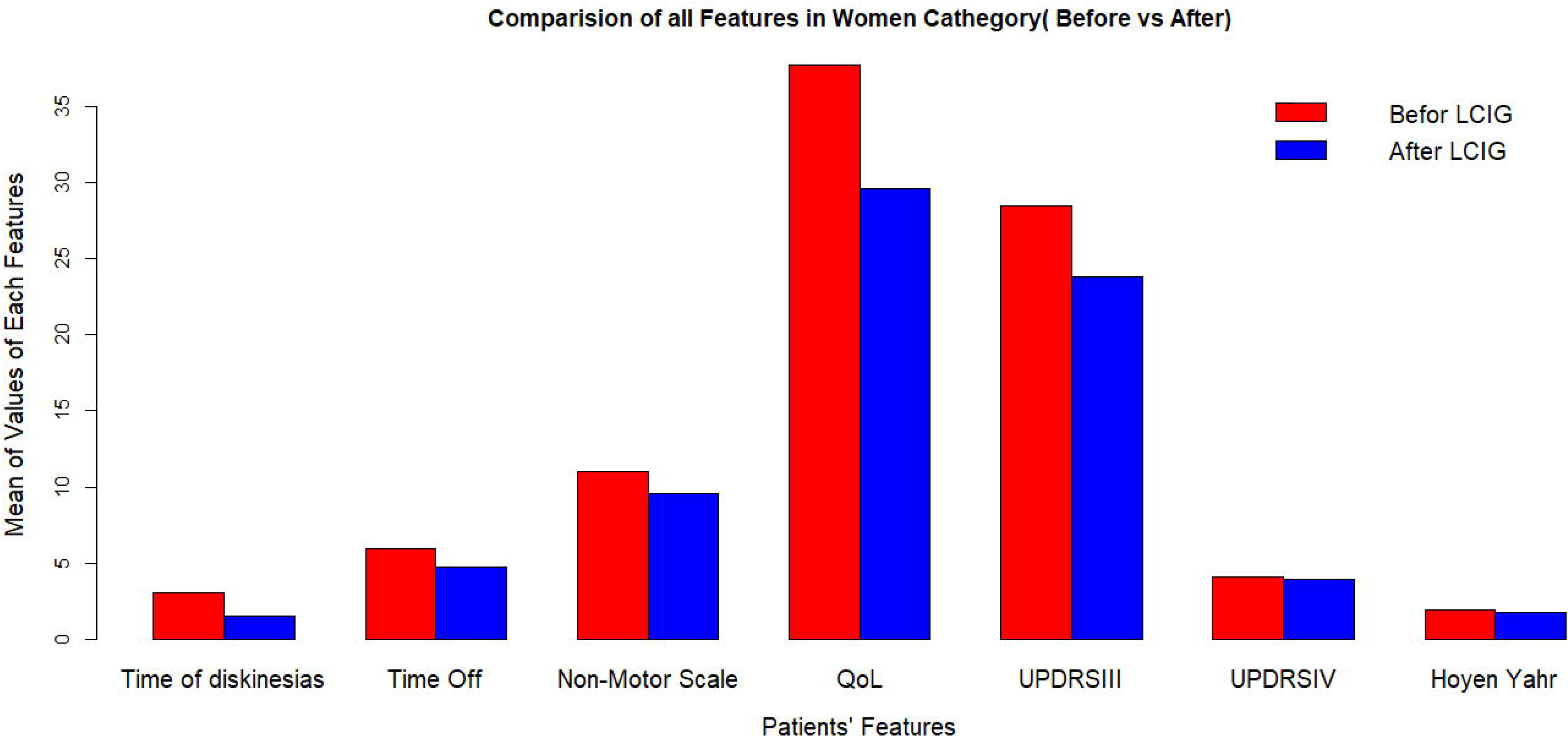
Clinical outcomes after LCIG implementation in women at 2-years follow-up

**Figure 2.**
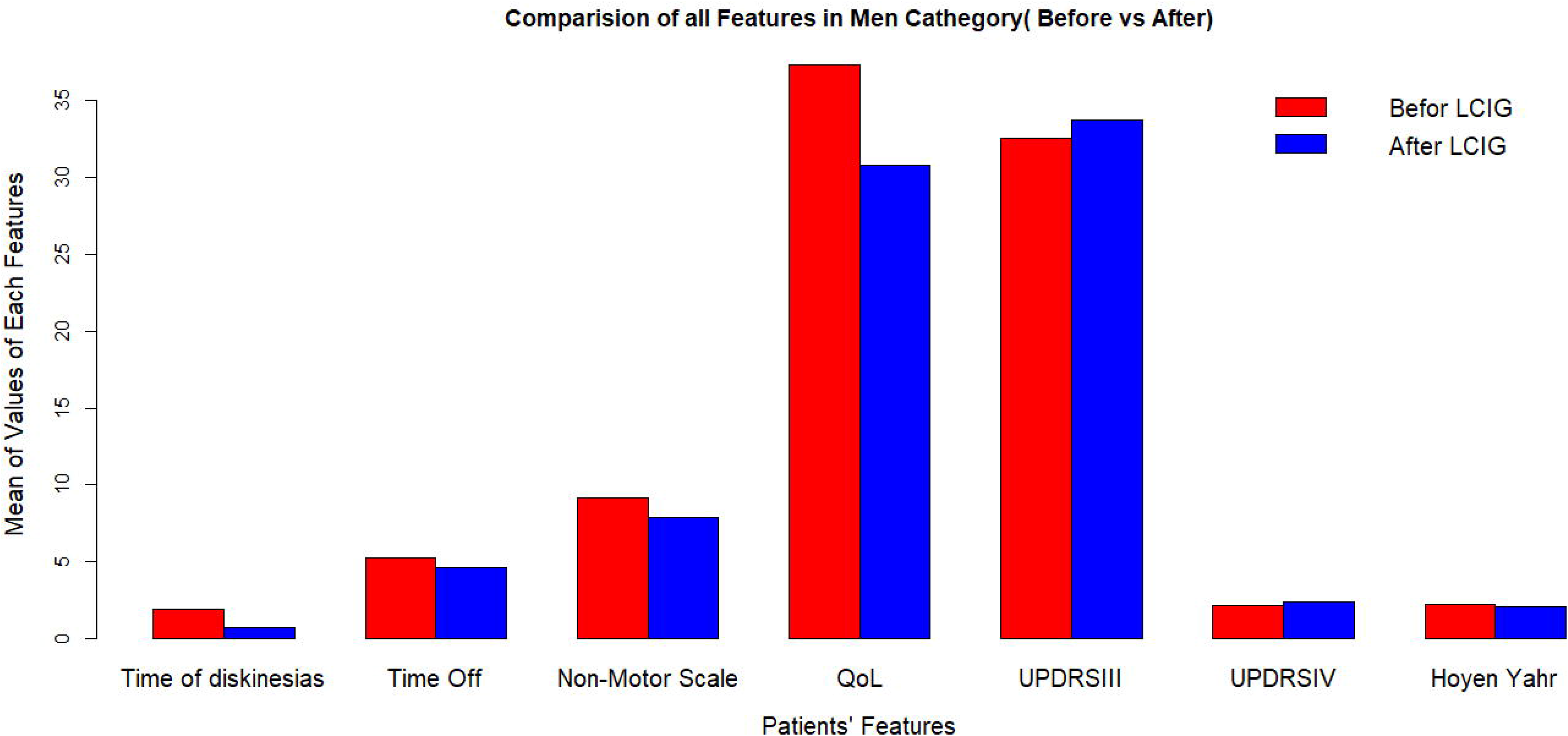
Clinical outcomes after LCIG initiation in men at 2-years follow-up

Shapiro-Wilk normality test confirms normal distribution of dyskinesia duration in women and men subgroups before and after LCIG treatment (p-value 0.448, 0.445, 0.049and 0.311respectively). F-test and box plot of data prove equality of variances of dyskinesia duration values for men and women before and after LCIG (p-values of the F-tests 0.653, 0.750, 0.951 and 0.943, respectively). Paired sample t test showed that dyskinesia duration is reduced for both women and men after LCIG treatment (p-values 1.151e -9 and 1.181e -8respectively).Two independent sample t test showed that the effect of LCIG treatment on dyskinesia duration is better on men compared to women (*P*= 0.002, Fig. 3).

**Figure 3.**
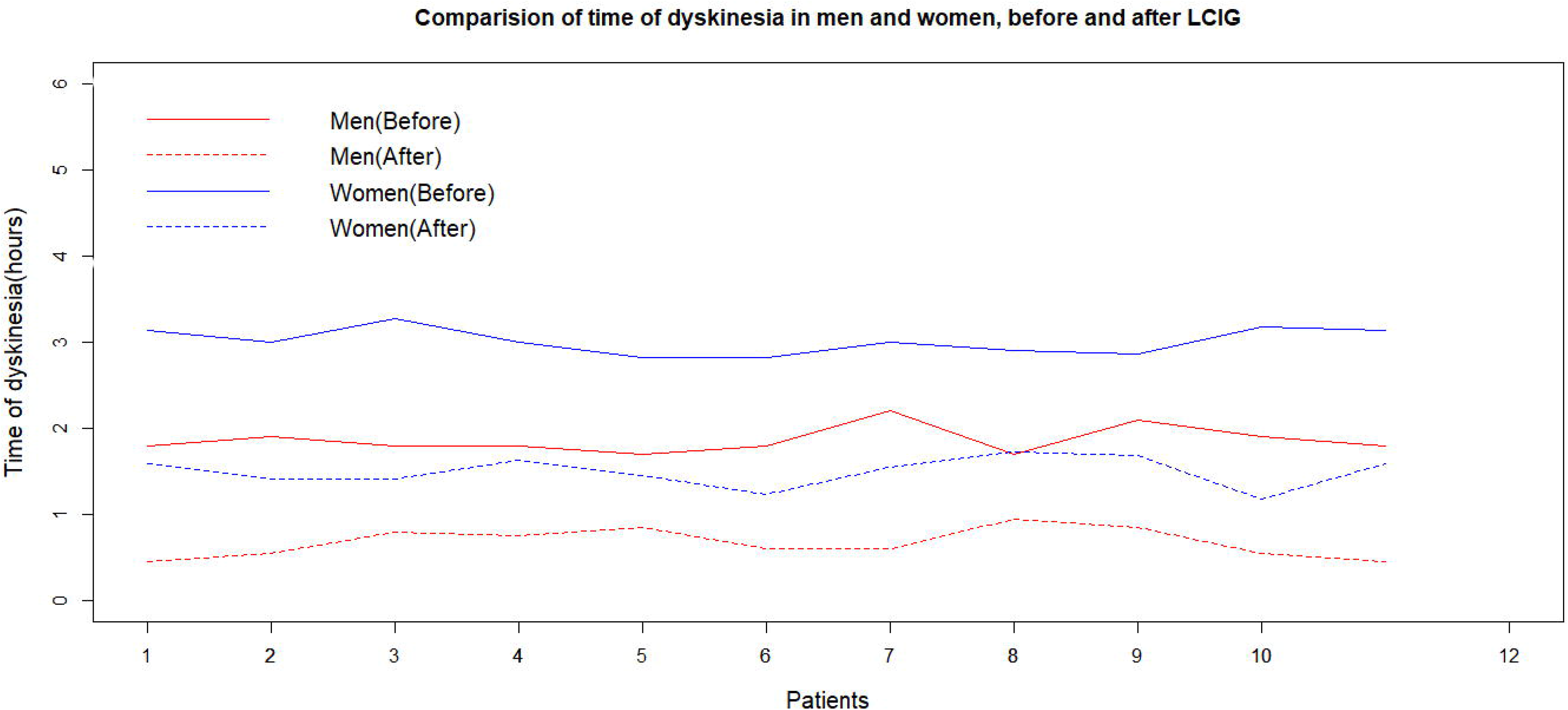
Effects of LCIG treatment on dyskinesia duration in men and women (before and 2 years after the initiation of LCIG treatment)

### Multivariate Linear Regression model

UPDRS-III (off) score was considered as the response variable, while NMSQ, PDQ-39, UPDRS-IV, HY scores, and dyskinesia duration at baseline as predictors. The slopes and intercept are 82, 0.20, -1.5, -4.39, 2.63 and 4.82. UPDRS-III score was decreased by 1.5 and 4.39 units per one unit of increase of the PDQ-39, UPDRS-IV indexes, respectively. UPDRS-III (off) score was also, increased 2.63 and 4.82 units per one unit of increase of the HY score and dyskinesia duration.

Dyskinesia duration is considered as the response variable, while NMSQ, PDQ-39, UPDRS-IV, HY and UPDRS-III (off) scores at baseline as predictors. The slopes and intercept are -0.45, -0.07, 0.09, 0.51, -0.81 and 0.009,respectively. Dyskinesia duration is increased by 0.51units per one unit of increase of the UPDRS-IV index. Dyskinesia duration is also decreased by 0.81unit per one unit of increase of the HY. The other features can be omitted from the linear model. It should be noted that this regression model investigated a linear relationship between dyskinesia duration as a response variable and NMSQ, PDQ-39, UPDRS-IV and HY at baseline as predictors, in which the most significant coefficient related to PDQ-39 and the other scores can be neglected. Dyskinesia duration increased one unit against half of a unit of PDQ-39 increment. In another linear regression model, PDQ-39 was considered as the dependent variable, while UPDRS-IV, UPDRS-III and HY as predictors. Therein, coefficient values are 0.467, -0.0476 and 2.489 respectively. PDQ-39 displays strong linear relation to HY scale. In another model, NMSQ score was supposed as the response variable for predictors UPDRS-III (off), UPDRS-IV and HY scores. Coefficients were 0.006, 0.786 and -0.699 respectively. This asserts that NMSQ score is dependent linearly to UPDRS-IV and HY scores.

### Mathematical Modeling

We present two types of models from two different approaches to predict the patients’ features with an acceptable horizon. First, among the time series models, we introduce the best model and estimate the model’s most accurate parameters for this purpose. AIC, Akaike information criterion will be used to find optimum parameters in the sequel. Second, a recurrent neural networks model will be used with a different approach to forecasting future values of these features and MSE, mean square error criterion will determine accuracy of the model. After preprocessing phase, since data are not equidistant, cubic spline interpolation has been used to convert this irregular time series to a regular one. The cycle of this time series is 31 days in a month and data are distributed across the spectrum. Fig.S1 is a plot of the produced scaled-regular time series with frequency 1 and without any outlier data. To omit possible outlier data. the lower and upper bounds are considered as 1.5 times IQR (interquartile rang) less than the first quartile and 1.5 times IQR greater than the second quartile, respectively.

### Time Series Models

In Fig.S1, the scaled-regularized time series is not stationary, there is an increasing trend and the seasonal effect still appears obvious, Augmented Dickey-Fuller test (ADF Test) and Kwiatkowski-Phillips-Schmidt-Shin test (KPSS test) confirm that time series is not stationary with *P*-values 0.9 and 0.1, respectively. So first of all the data should be detrend and seasonality be removed. Therefore 1 lag and 1 time differentiation is taken to decrease ADF test p-value to 0.01, this proves stationary status is achieved and also the plot of obtained time series confirms that produced signal is stationary (Fig.S1). The shape of the plot suggests ARMA model (combine two models, Auto Regression (AR) and Moving Average (MA)) is probably better than a single-cyclesinusoidal. We fit and obtain the optimum parameter(s) estimates for AR, MA and SARIMA models and choose the best model’s parameters based on minimum Akaike information criterion, AIC. In AR models, p=1 is the best estimate, which P: indicate number of autoregressive terms (lags of the stationarized series). For MA models, q=1 is the best estimate, where q Trend moving average order. In all SARIMA model parameters P=1, q=0 and d=1 are the best main parameters and P=2, q=0 and d=1 are the best parameters for seasonality and also *S*=10 is the time span of repeating seasonal pattern. AIC criterion for these three models is -80, -28 and -133 respectively, then the best model is MA model. It is worthwhile to note that the exponential models are ignored because in this project it was the worst in compared with SARIMA models. Also, exponential time series model with AIC -18 is better than all ARMA models and at last all of these models fail to be fit well as the LSTM model and this is significant for long horizon of prediction (Fig. 4).

**Figure 4.**
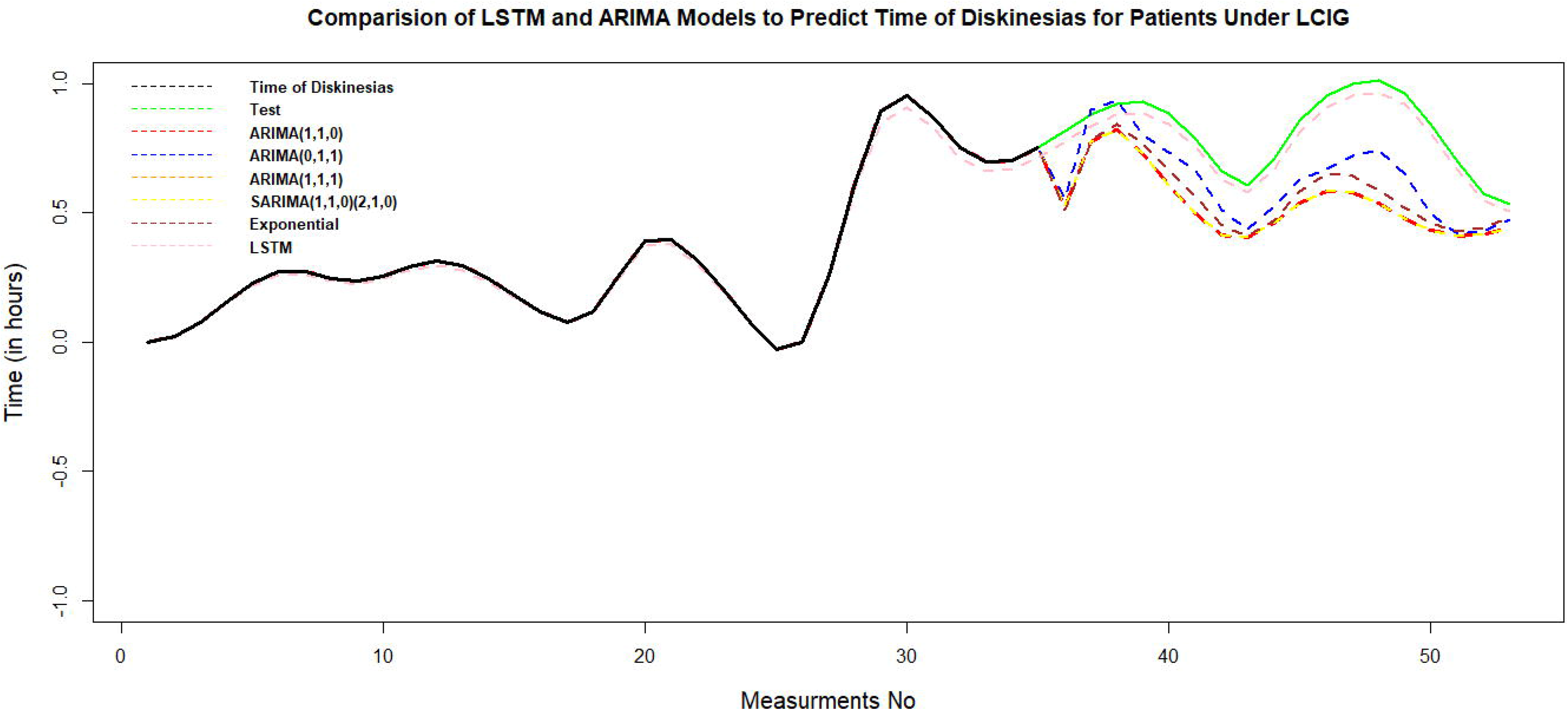
Comparison of LSTM and ARIMA Models to predict Dyskinesia duration for Patients Under LCIG

### LSTM Model

After preparing the data set and removing outlier data by a suitable wavelet transform, an LSTM algorithm has been trained by 80% of data set, and it has been tested by the second part of that. In Fig. 4 green and pink curvatures display the test and predicted data of dyskinesia duration in using LSTM method; Although it can be seen that pink and green dashed lines are coincides well, to measure accuracy of the LSTM model the error value is computed by mean square error, MSE=0.0069 shows the accuracy of this model.

## Discussion

This study presents simple visualization tools that show trends evolving or repeating over time to advanced Μl to discover the specific structure of time series related to the PD patients’ data set. Our harmonic analysis suggests that dyskinesia duration can be significantly improved under LCIG therapy. The UPDRS-III (off) score variation under LCIG therapy is sex dependent. Different time series models are fitted to predict time of dyskinesia after LCIG therapy. Among these models, exponential model is the best one with AIC criterion -18 and MSE=0.02. However, the traditional times series models fail to predict values of PD patients’ features in a long horizon of time. An RNN model adjusted to predict these features. To our knowledge, this is the first attempt to predict motor and non-motor features of patients with advanced PD under LCIG using a ML such RNN-LSTM model. MSE=0.008 showed high accuracy to predict future PD features in test part of data.

A novel finding of this study was the better effect of the LCIG on time of dyskinesia in men than in women. In line with previous studies, women are more likely than men to develop dyskinesia before the LCIG treatment^16–19^. Although clinical trials with LCIG were successful in reducing motor symptoms including dyskinesia, sex differences were not fully assessed^21–28^. The etiology of sex effect is not clear. It has been suggested that hormonal status may modify sensitivity to levodopa^16^, and that lower body weight may lead to greater levodopa exposure ^17^. Higher LEDD (≥ 2000 mg/day) was required in men than in women^20^. Higher discontinuation rate due to non-procedure or device-associated adverse effects was noted in the ≥ 2000 mg/day group ^20^. More future studies will be useful in decision making on LCIG therapy for individual patients.

In the present study, LCIG treatment significantly improved the QoL assessed by PDQ-39, in agreement with previous reports^21–23, 27^. Specifically, UPDRS-III was decreased by 1.5 and 4.39 units per one unit of increase of the PDQ-39, similar to the findings of other studies ^21, 26, 27^. We observed UPDRS-IV, UPDRS-III and HY scores as predictors for the PDQ-39 score. HYshowed a strong association with PDQ-39. A possible explanation could be that poor QoL reflects greater severity of the disease later in the disease course.

Importantly, UPDRS-III (off), UPDRS-IV and HY scores were predictors for the total score of the NMSQ. However, the HY scale does not necessarily reflect therapy=:Jrelated improvements in NMS, which are not well captured by this staging system.On the other hand, the burden of NMS at baseline predicts improvement in QoL in patients treated with LCIG ^28^. However, these studies were single-centered and not longitudinal including non naïve PD patients.

Our results have several implications. Using LSTM, time series forecasting models can predict treatment outcomes based on previous, sequential data. This provides greater accuracy for ML methods LSTM-based recurrent neural networks are probably the newest powerful approach to learning from sequential data. The LSTM cell adds long-term memory in an even more performant way because it allows even more parameters to be learned. This makes it the most powerful recurrent neural network to do forecasting, especially when you have a longer-term trend in your data Previous studies have employed such approaches and used clinical, genetic, imaging, and cerebrospinal fluid markers to predict the initiation of symptomatic therapy and motor progression based on UPDRSIII. This research is a small, but determined, step in attaining a better view and knowledge regarding the more advanced means and their usage in ML applications.

Our study also has some limitations. PD is a non homogeneous disease with many subtypes. We did not explore subtypes of PD and whether our artificial neural network equally well with all subtypes. Another limitation of this study is that both the progression analysis and preclinical diagnosis were validated in a small number of participants. Larger studies are required to further confirm these results. Further, while we have tested the model across institutions and using independent datasets, future studies should expand the diversity of datasets and institutions. However, the mechanisms that lead to the development and progression of motor and non-motor symptoms in advanced PD are incompletely understood and need further analysis.

Nonetheless, the strengths of our study include a very thorough assessment, a longitudinal follow-up design, and the extensive clinical and demographic information recorded in ten time points. By tracking the progression of early, initially untreated PD and including only Part III scores in the *off*=:Jmedication state, potential confounding from medication effects at baseline assessments has been reduced. In the new era of artificial intelligence, our RNN-LSTM model could potentially be used as an additional tool in health decision-making, to predict disease progression in PD patients under LCIG therapy.

## Data Availability

All data produced in the present study are available upon reasonable request to the authors

## Acknowledgements

none

## Supplementary material

**Figure S1.** Irregular Time Series and Regularized Time Series Without Outliers, Using Qubic Spline.

Ethical and Regulatory Aspects

This study was conducted in accordance with the Declaration of Helsinki and the current European Data Protection Regulation. Written informed consent was obtained from all participants. We confirm that we have read the Journal’s position on issues involved in ethical publication and affirm that this work is consistent with those guidelines. The authors take full responsibility for the integrity of the data and the accuracy of the data analysis.

Data Availability Statement

The data supporting the findings of this study are available upon reasonable request from any qualified investigator.

Authors’ Roles:

(1) Research project: A. Conception, B. Organization, C. Execution; (2) Statistical Analysis: A. Design, B. Execution, C. Review and Critique; (3) Manuscript: A. Writing of the first draft, B. Review and Critique.

Anastasia Bougea: 1A,1B,1C,3A,3B

Tajedin Derikvand: 2A,2B, 2C,3B

Efthymia Efthymiopoulou 1A,1B,2C,3B

Efthalia Angelopoulou 1B,2C,3B

Financial Disclosures of all authors (for the preceding 12 months)

There are no specific disclosures related to the current work.

Anastasia Bougea is an investigator in studies funded by AbbVie; the Michael J. Fox Foundation for Parkinson Research; the “ALAMEDA” study (H2020-EU, Grant Agreement 101017558). She has received travel grant, or speaker honoraria from AbbVie.

Tajedin Derikvand: has no disclosures.

Efthymia Efthymiopoulou is an employee of AbbVie and may hold AbbVie stock and/or stock options.

Efthalia Angelopoulou has no disclosures.

## References

1. Poewe, W., Seppi K, Tanner CM, et al. Parkinson disease. Nat Rev Dis Primers 2017; 3:17013.

2. Chaudhuri KR, Schapira AH. Non-motor symptoms of Parkinson’s disease: dopaminergic pathophysiology and treatment. Lancet Neurol 2009;8: 464–74.

3. Fasano A, Fung VSC, Lopiano L, et al. Characterizing advanced Parkinson’s disease: OBSERVE-PD observational study results of 2615 patients. BMC Neurol 2019; 19:50.

4. Antonini A, Odin P, Pahwa R, et al. The Long-Term Impact of Levodopa/Carbidopa Intestinal Gel on ’Off’-time in Patients with Advanced Parkinson’s Disease: A Systematic Review. Adv Ther 2021; 38(6):2854–2890.

5. Emrani S, McGuirk A, Xiao W. Prognosis and diagnosis of Parkinson’s disease using multi-task learning. KDD 2017; 1457–66.

6. Shamir RR, Dolber T, Noecker AM, Walter BL, McIntyre CC. Machine Learning Approach to Optimizing Combined Stimulation and Medication Therapies for Parkinson’s Disease. Brain Stimul 2015; 8(6):1025–32.

7. Efthymiopoulou E, Antonoglou A, Loupo B, Bougea A. Determination of the motor status of patients with advanced Parkinson’s disease under levodopa-carbidopa intestinal gel using a machine learning model. Acta Neurol Belg. 2023;123(2):565–570.

8. Byeon H. Is the Random Forest Algorithm Suitable for Predicting Parkinson’s Disease with Mild Cognitive Impairment out of Parkinson’s Disease with Normal Cognition? Int J Environ Res Public Health 2020;17(7):2594.

9. Siami-Namini S, Tavakoli N, Siami Namin A. A Comparison of ARIMA and LSTM in Forecasting Time Series,17th IEEE International Conference on Machine Learning and Applications (ICMLA) 2018;1394–1401.

10. Houdt GV, Mosquera C, Nápoles G. A review on the long short-term memory model. Artificial Intelligence Review 2020; 1–27.

11. Fahn S, Elton R, and Members of the UPDRS Development Committee Unified Parkinson’s Disesae Rating Scale. In: Fahn S, C Marsden, D Calne, M Goldstein (ed) Recent development in Parkinson’s disease. Florhan Park: Macmillan Health Care Information, 1987 pp 153–64.

12. Chaudhuri KR, Martinez-Martin P, Schapira AH, et al. International multicenter pilot study of the first comprehensive self-completed nonmotor symptoms questionnaire for Parkinson’s disease: the NMSQuest study. Mov Disord 2006;21(7):916–23.

13. Yesavage JA, Brink TL, Rose TL, Lum O, Huang V, Adey M, Leirer VO. Development and validation of a geriatric depression screening scale: a preliminary report. J Psychiatr Res 1983; 17(1):37–49

14. Hoehn MM, Yahr MD. Parkinsonism: onset, progression and mortality. Neurology 1967; 17:427–442.

15. Jenkinson C, Fitzpatrick R, Peto V, Greenhall R, Hyman N. The Parkinson’s Disease Questionnaire (PDQ-39): development and validation of a Parkinson’s disease Summary Index Score. Age Ageing 1997;26: 353–357

16. Fabbri M, Zibetti M, Calandra-Buonaura G, et al. Levodopa/Carbidopa Intestinal Gel Long-Term Outcome in Parkinson’s Disease: Focus on Dyskinesia. Mov Disord Clin Pract. 2020;7(8):930–939.

17. Zappia M, Annesi G, Nicoletti G et al. Sex differences in clinical and genetic determinants of levodopa peak-dose dyskinesias in Parkinson disease: an exploratory study. Arch Neurol 2005; 62(4):601–605

18. Russillo MC, Andreozzi V, Erro R, Picillo M, Amboni M, Cuoco S, Barone P, Pellecchia MT. Sex Differences in Parkinson’s Disease: From Bench to Bedside. Brain Sci. 2022; 13;12(7):917.

19. Santos-García D, Laguna A, Hernández-Vara J, et al., On Behalf Of The Coppadis Study Group. Sex Differences in Motor and Non-Motor Symptoms among Spanish Patients with Parkinson’s Disease. J Clin Med. 2023;12(4):1329.

20. Zadikoff C, Poewe W, Boyd JT, Bergmann L, Ijacu H, Kukreja P, Robieson WZ, Benesh J, Antonini A. Safety of Levodopa-Carbidopa Intestinal Gel Treatment in Patients with Advanced Parkinson’s Disease Receiving ≥2000=:Jmg Daily Dose of Levodopa. Parkinsons Dis. 2020:9716317.

21. Fernandez HH, Standaert DG, Hauser RA, et al. Levodopa carbidopa intestinal gel in advanced Parkinson’s disease: final 12-month, open-label results. Mov Disord 2015;30(4):500–509.

22. Olanow CW, Kieburtz K, Odin P, et al. Continuous intrajejunal infusion of levodopa-carbidopa intestinal gel for patients with advanced Parkinson’s disease: a randomised, controlled, double blind, double-dummy study. Lancet Neurol 2014;13(2):141–149.

23. Slevin JT, Fernandez HH, Zadikoff C, et al. Long-term safety and maintenance of efficacy of levodopa-carbidopa intestinal gel: an open-label extension of the double-blind pivotal study in advanced Parkinson’s disease patients. J Parkinsons Dis 2015;5(1):165–174.

24. Antonini A, Yegin A, Preda C, et al. Global long-term study on motor and non-motor symptoms and safety of levodopa-carbidopa intestinal gel in routine care of advanced Parkinson’s disease patients; 12-month interim outcomes. Parkinsonism Relat Disord 2015;21(3):231–235.

25. Freire-Alvarez E, Kurča E, Lopez Manzanares L, Pekkonen E, Spanaki C, Vanni P, Liu Y, Sánchez-Soliño O, Barbato LM. Levodopa-Carbidopa Intestinal Gel Reduces Dyskinesia in Parkinson’s Disease in a Randomized Trial. Mov Disord. 2021;36(11):2615–2623.

26. Kovács N, Bergmann L, Anca-Herschkovitsch M, Cubo E, Davis TL, Iansek R, Siddiqui MS, Simu M, Standaert DG, Chaudhuri KR, Bourgeois P, Gao T, Kukreja P, Pontieri FE, Aldred J. Outcomes Impacting Quality of Life in Advanced Parkinson’s Disease Patients Treated with Levodopa-Carbidopa Intestinal Gel. J Parkinsons Dis. 2022;12(3):917–926.

27. Standaert DG, Rodriguez RL, Slevin JT, Lobatz M, Eaton S, Chatamra K, Facheris MF, Hall C, Sail K, Jalundhwala YJ, Benesh J. Effect of levodopa-carbidopa intestinal gel on non-motor symptoms in patients with advanced Parkinson’s disease. Mov Disord Clin Pract. 2017;4: 829–837.

28. Chaudhuri KR, Antonini A, Robieson WZ, Sanchez Solino O, Bergmann L, Poewe W. Burden of non-motor symptoms in Parkinson’s disease patients predicts improvement in quality of life during treatment with levodopa-carbidopa intestinal gel. Eur J Neurol. 2019; 26:581–e543.

