## Supplementary figures and images for "Artificial neural network predicts sex differences of patients with advanced Parkinson’s disease under Levodopa-Carbidopa Intestinal gel"

### Supplemental Figure 1

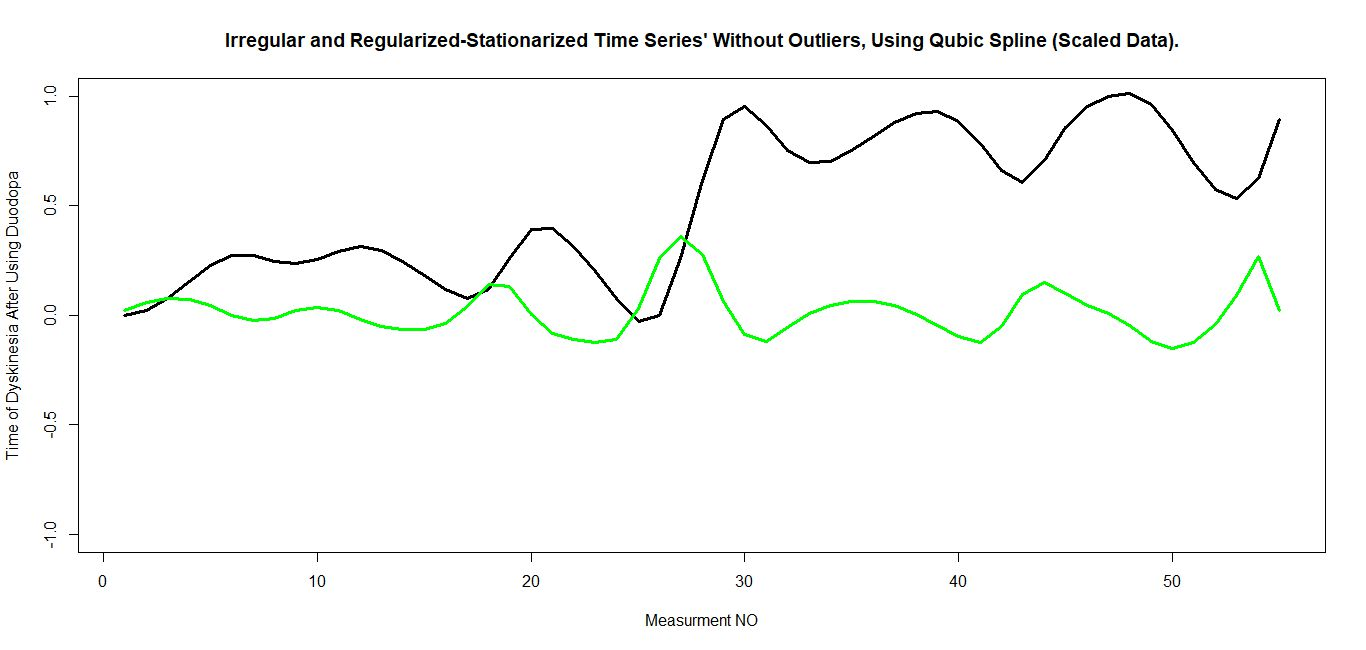
